# Monitoring for outbreak associated excess mortality in an African city: Detection limits in Antananarivo, Madagascar

**DOI:** 10.1101/2020.10.18.20214411

**Authors:** Fidisoa Rasambainarivo, Anjarasoa Rasoanomenjanahary, Joelinotahiana Hasina Rabarison, Tanjona Ramiadantsoa, Rila Ratovoson, Rindra Randremanana, Santatriniaina Randrianarisoa, Malavika Rajeev, Bruno Masquelier, Jean Michel Heraud, C. Jessica E. Metcalf, Benjamin L. Rice

**Author notes:** These authors contributed equally to this work (Fidisoa Rasambainarivo, Anjarasoa Rasoanomenjanahary, Joelinotahiana Hasina Rabarison). These authors supervised this work equally (Bruno Masquelier, Jean Michel Heraud, C. Jessica E. Metcalf, Benjamin L. Rice).

## Abstract

Quantitative estimates of the impact of infectious disease outbreaks are required to develop measured policy responses. In many low- and middle-income countries, inadequate surveillance and incompleteness of death registration are important barriers. Here, we characterize how large an impact on mortality would have to be to be detectable using the uniquely detailed mortality notification data from the city of Antananarivo in Madagascar, with application to a recent measles outbreak. The weekly mortality rate of children during the 2018-2019 measles outbreak was 154% above the expected value at its peak, and the signal can be detected earlier in children than in the general population. This approach to detecting anomalies from expected baseline mortality allows us to delineate the prevalence of COVID-19 at which excess mortality would be detectable with the existing death notification system in the capital of Madagascar. Given current age-specific estimates of the COVID-19 fatality ratio and the age structure of the population in Antananarivo, we estimate that as few as 11 deaths per week in the 60-70 years age group (corresponding to an infection rate of approximately 1%) would detectably exceed the baseline. Data from 2020 will undergo necessary processing and quality control in the coming months. Our results provide a baseline for interpreting this information.

## Introduction

During an infectious disease outbreak, the reported number of deaths may underestimate the true mortality caused by the disease. Both changing case definitions (Tsang et al., 2020), and a lack of access to testing can bias reported cause-specific mortality counts, as repeatedly noted during the COVID-19 pandemic crisis (Kiang et al., 2020). One approach to evaluating the magnitude of the bias in such burden estimates is by estimating the deviation of total number of deaths from the expected baseline levels of mortality from previous years, a measure called “excess deaths”. By avoiding the issue of misclassification of deaths, this approach provides an upper bound on excess mortality directly due to the infection. Since both direct and indirect mortality (e.g., resulting from individuals being denied access to routine care) are included, the true direct burden may be lower than this estimate, but the combined quantity provides important policy information as to the overall severity and impact of the health crisis. This information may help target interventions, compare between subgroups of a population, locations or periods and draw lessons for management of future public health crisis (Dowd et al., 2020; Leon et al., 2020).

In the case of COVID-19, evidence from high-income settings indicates that the excess may amount to up to 700% above the expected mortalities of a locality, primarily concentrated in older age groups (Weinberger et al., 2020). The rapid acceleration of the infection fatality ratio of SARS-CoV-2 with age has led to the suggestion that the mortality impact of the pandemic will be lower in low- and middle-income countries (LMIC) with younger age structures (Dowd et al., 2020; Salje et al., 2020; Verity et al., 2020). Nevertheless, the impact of comorbidities and disruption of other aspects of the health system might still drive considerable excess mortality (Rice et al., 2020). However, to date, analyses of excess mortality have been largely confined to high income, western countries and cities (Marcon et al., 2020; Michelozzi et al., 2020; Signorelli et al., 2020; Sinnathamby et al., 2020; Weinberger et al., 2020; Woolf et al., 2020) in part because accurate data on mortality from previous years is required: few countries in lower income settings have statistical agencies and reporting mechanisms that have the capacity and infrastructure to report the number of people that died on a weekly or on a day-to-day basis.

While death registration systems are often deficient at the national level, some urban centers maintain complete and up-to-date registers. In Antananarivo, the capital city of Madagascar, detailed mortality data maintained since 1976 have provided a unique window onto questions ranging from the drivers of the epidemiological transition in low income settings to changes in seasonality in mortality across four decades (Masquelier et al., 2019; Schlüter et al., 2020). This unique data opens the way to detecting disease driven-anomalies in mortality data in a lower income setting as was shown during the 2009 Influenza (AH1N1) virus pandemic (Rajatonirina et al., 2013). However, the higher incidence of infectious diseases in such settings also drives strong background fluctuations in mortality (seasonal and often age specific) that must be accounted for, and might impede detection of mortality anomalies. For instance, Schlüter (Schlüter et al., 2020) et al showed that in Antananarivo, deaths in adults >65 years old peak during the austral winter months (June-September) while that of children peak in December -January. Therefore, an unexpected increase in mortality of elder individuals, concurrent with the decrease in mortalities of children or vice-versa may be inapparent or appear to be of a lesser extent when considering the entire population.

Here, we apply classical statistical tools to this unique database to quantify excess mortality during a severe measles outbreak that occurred in 2018-2019. We use this as a foundation to explore the detection limits of the system during the COVID-19 pandemic by leveraging what is known about the mortality profile of COVID-19 in order to delineate the degree to which excess deaths would be detectable with the existing death notification system. Because death notification records for 2020 were unavailable to us at the time of writing, our analysis only lays the foundations for a retrospective analysis of the impact of the pandemic in a LMIC setting.

## Methods

Antananarivo-Renivohitra, the capital district of Madagascar, has a population estimated at 1 275 207 in 2018 (RGPH-3, 2018), of which 32.16% are under 15 years old and only 5.5% are over 60 years old. All residents’ death records which include the date of birth, date of death, age and cause of death ascertained by a physician, covering the period from 2010 to 2019 were transcribed from registers maintained by the Municipal Office of Hygiene (BMH) of the city of Antananarivo.

### Estimating the measles outbreak excess mortality

In order to establish the baseline death counts in Antananarivo in the absence of the measles outbreak, we fit an autoregressive quasi poisson and an autoregressive negative binomial regression model adapted for time series analysis (Liboschik et al., 2017) to the weekly counts of deaths between January 01, 2010 and the day preceding the detection of the first detected case of measles in the city (August 31st, 2018), separating the data into epidemiologically relevant age groups (0-14,15-59 and 60+ years old) based on known features of the epidemiology of measles. Short range serial dependence is captured by regressing on the previous weeks and the seasonality is captured by regressing on the conditional mean of 52 weeks (1 year) back in time. We also include a deterministic covariate to account for a yearly trend. For each age group, we compare the fit of the Poisson model with that of the negative binomial via the proper scoring rules and marginal calibration plots. We then fit different models in which we vary the number of weeks capturing the orders of autoregressive terms and include or exclude the seasonal and trend components. The best performing model was selected by minimizing the Akaike Information Criterion (AIC). This model was then projected forward using one-week ahead projections for the duration of the measles outbreak (39 weeks) to forecast the expected number of deaths (and 95% Confidence Interval) in the absence of the disease. Excess deaths are detected when observed mortality is above the upper bound of 95% CI. To calculate the total outbreak-attributable number of excess deaths, we subtracted the weekly model-predicted values from the observed number of deaths reported and summed across the duration of the outbreak.

### Estimating expected mortality in 2020

In order to estimate the baseline death counts expected for the year 2020 for each age category, we fit similar models to the time series spanning from January 1st, 2010 to December 31st, 2019 disaggregated into 10-year bands between 0-9 years and 70-79 and a further age band for individuals of 80 years and older. Using the best model, we forecast the expected number of deaths per week for the next 52 weeks which can be used to assess the burden of COVID-19 in Antananarivo for the year 2020.

Using published infection fatality ratio from studies in France and China ^5,6^ and the age-structure of the population of Antananarivo from the 2018 census, we estimate the expected number of deaths per age category at a given prevalence of infection. The expected number of deaths per age category is given by the equation:

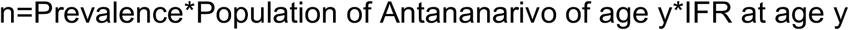

Finally, we estimate the prevalence of infections that will lead to mortality rates that are significantly above the expected baseline.

Statistical analyses were performed in R and using the package ‘tscount’ (autoregressive negative binomial and poisson models) and mgcv.

## Results

Between January 1st 2010 and December 31st 2019, 68 820 deaths were reported from residents of the Antananarivo Renivohitra district. Of these, 950 were of children (0-14 years old) during the measles outbreak spanning from September 01, 2018 to May 22, 2019.

Our results indicate that the observed number of deaths recorded during the measles outbreak epidemic was significantly higher than expected, particularly in children (figure 1). Death counts of all ages and from all causes were significantly above the expected values during 10 of the 39 weeks of the outbreak starting on 2018-12-16. In contrast, when considering the younger age group (0-14 years old), most vulnerable to the disease, excess mortalities were detected as early as 2018-10-28 and were significantly higher than expected during 16 weeks with 39 deaths per week on average compared to the 18 deaths expected in each of those weeks. At its peak, the mortality rate of children during the measles outbreak was 161% above the expected value (47 deaths/ week). The total excess deaths for individuals younger than 15 years old during the measles outbreak period is estimated at 319. Of those, only 122 report measles as a cause of death. In adults (between 15-59 years old), mortality exceeded the baseline for 6 weeks starting on 2018-12-16 in the course of which, we estimate the excess mortality to be at 247 deaths but only 3 adult deaths recorded during the outbreak period were attributed to measles. No excess deaths were detected in elderly residents.

**Figure 1:**
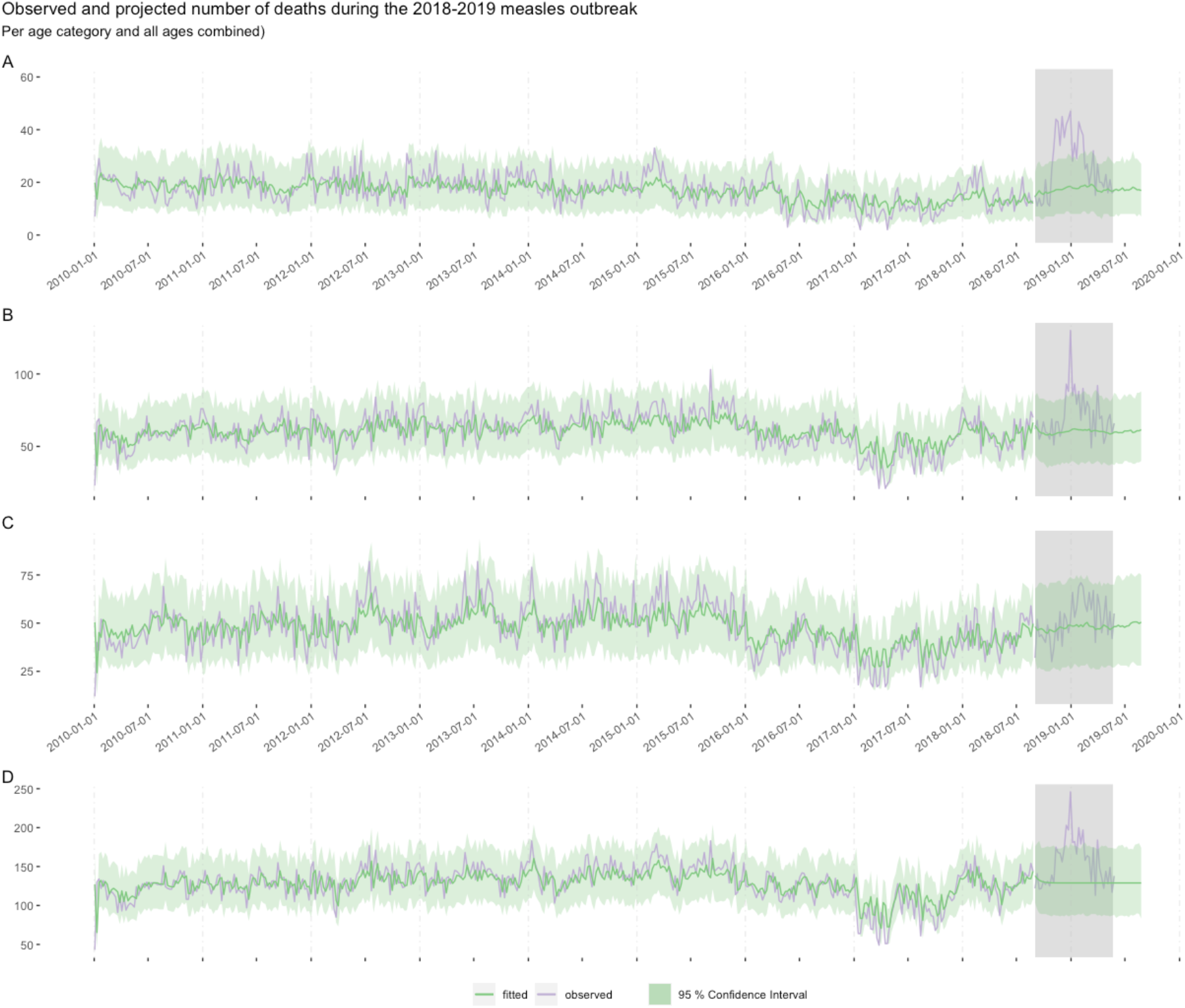
Time series of the observed and projected weekly death counts of the residents of Antananarivo from 2010-01-01 to 2019-05-22 per age category (A: <15 years old, B: 15-60 years old and C: >60 years old) and the entire population (D). Grey shaded areas indicate the period of the measles outbreak.

This unique dataset allows us to establish the baseline of death counts for the population of Antananarivo in the absence of COVID-19 which is necessary to evaluate the burden of the pandemic on the population. Based on the model fitted to the data until December 2019, we estimated that the expected weekly number of deaths in adults of more than 60 years old, is 51 deaths per week with an upper bound of the 95% confidence interval of 79 deaths per week. Therefore, an excess of 28 deaths per week or more of the elderly residents would be significantly higher than expected and detectable with the existing death notification system in Antananarivo, Madagascar.

Given the age structure of the population of Antananarivo (figure 2) and published estimates of the IFR (Salje et al., 2020; Verity et al., 2020), we computed the prevalence of SARS-CoV-2 infections that will directly lead to a detectable excess of mortality for each age group (figure 2).

**Figure 2:**
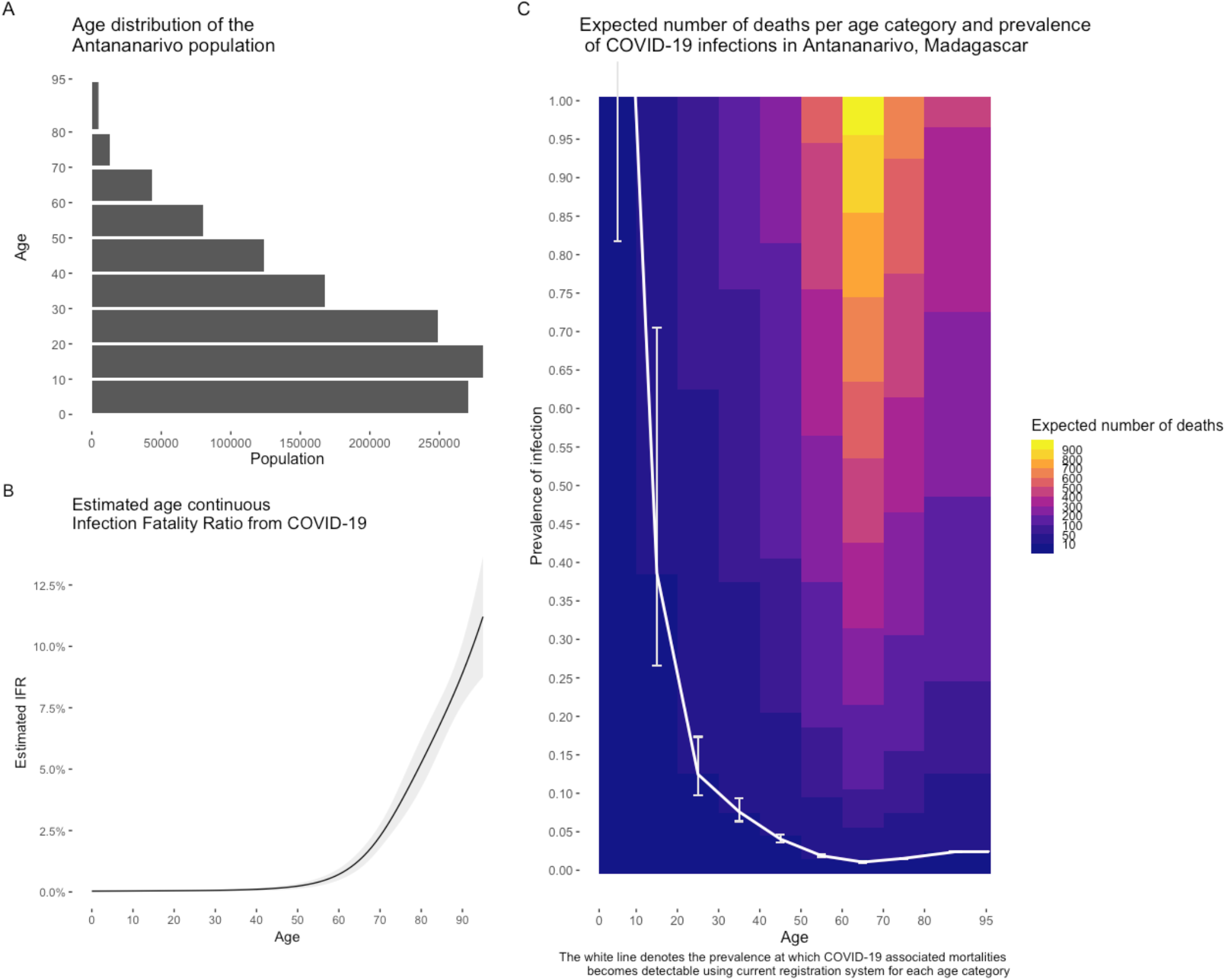
Expected number of deaths per age group as a function of the prevalence of infection and the age structure of the population in Antananarivo, Madagascar given published estimates of IFR. White line indicates the prevalence of COVID-19 at which the burden of the disease would be detectable (higher than the difference between upper bound confidence interval and model-based estimates).

## Discussion

During outbreaks of deadly diseases, an increase in mortality is anticipated as a consequence of the disease itself and the potential disruption of the healthcare system. Here, we demonstrate detectability of the excess mortality from a measles outbreak in the capital city of Madagascar and establish a baseline to assess the impact of the COVID-19 pandemic in the population. First, our results show that the current death notification system can detect outbreak-driven anomalies in mortality and illustrate how the signal may be detected earlier in age stratified time series. Our results also suggest that the measles outbreak had a higher burden on the residents of Antananarivo than previously reported. Drivers of this excess mortality could include misclassification of the cause of death, indirect impacts of measles via disruption in the health provision or immunosuppression leaving individuals vulnerable to other infections (Mina et al., 2015).

Second, this unique dataset allows us to establish the baseline of mortalities for the population of Antananarivo in the absence of COVID-19 which is necessary to evaluate the burden of the pandemic on the population, provided that there are no massive in- out- migrations flows, due to the lockdown measures, and that the death reporting systems remains functional during the pandemic. We estimate that, given what is known to date about the infection fatality rate over age, it is virtually impossible for children’s mortality to rise above the expected baseline (17 deaths per week 95%CI: 6-30) as a direct consequence of infection by SARS-CoV-2. In fact, given the current IFR estimates for this age group (0.0026% CI:0.0002-0.002%-Figure 2), even if all of the estimated 247 000 children of less than 10 years old were infected, the excess mortality would not exceed the baseline death counts. Therefore, if any excess deaths are reported in this age category for the year 2020, they are likely to result from indirect effects of the pandemic on child mortality (Roberton et al., 2020). In contrast, only 1% of the 47 575 individuals between 60 and 70 years old, would need to be infected with SARS CoV-2 in any given week to be subsequently detectable using the current death notification system.

We base our estimates of the detectable prevalence of SARS-CoV-2 infection from the death notification system in Antananarivo using currently available detailed estimates of the age-specific IFR, which all emerge from higher income countries, with specific rates of non-communicable diseases (Italy, France) (Salje et al., 2020; Verity et al., 2020). The higher rates of noncommunicable diseases in younger ages in many cities of LMIC compared to other urban centers may shift the IFR profile (Rice et al., 2020), increasing the burden of mortality following infection in these settings. The result would be that lower prevalence would be detectable through this death notification system, but also that the burden of the pandemic would be greater. The pattern of IFR over age for LMICs, including in Madagascar is an important knowledge gap. As many lines of evidence suggest that demographic, but also economic and social characteristics as well as health care provision all play important roles, we also expect considerable sub-national variation in LMICs, including in other urban centers and rural regions of Madagascar. The dataset collected by the BMH offers a unique opportunity to provide a first data-point to better understand this diversity, as well as an assessment of the effects of a public health crisis on the residents of Antananarivo, the capital city of Madagascar, with important implications for identifying strategies to mitigate the COVID-19 pandemic.

## Data Availability

Code and data are available in a publicly available online repository: https://github.com/fidyras/mortality

## Appendix 1 Model selection and best performing models

### Model selection

|  | All ages | Young | Adults | Elderly |
| --- | --- | --- | --- | --- |
| past_obs=1 (beta 1) | **3887.74** | 2740.45 | 3393.16 | 3353.00 |
| past_obs=2 (beta 2) | 3952.76 | 2769.64 | 3409.21 | 3377.43 |
| past_obs=3 (beta 3) | 3928.83 | 2783.13 | 3393.55 | 3378.44 |
| past_obs=4 (beta 4) | 3973.64 | 2803.50 | 3432.81 | 3393.19 |
| past_obs=1 (beta 1) + past_mean=52 (alpha 52) | 3889.09 | 2729.71 | **3391.93** | 3350.13 |
| past_obs=1 (beta 1) + linear trend | 3889.18 | 2713.55 | 3393.59 | 3349.93 |
| past_obs=1 (beta 1)+ past_mean=52 (alpha 52) + linear trend | 3890.69 | **2711.30** | 3392.81 | **3347.12** |

Model weekly death counts in children (0-14 years old) of Antananarivo Coefficients:
|  | Estimate | Std.Error | CI(lower) | CI(upper) |
| --- | --- | --- | --- | --- |
| (Intercept) | 0.9748 | 0.28471 | 0.4168 | 1.53286 |
| **Beta 1** | **0.4157** | **0.04361** | **0.3302** | **0.50118** |
| **Alpha 52** | **0.2586** | **0.09744** | **0.0677** | **0.44962** |
| **Linear Trend** | **-0.0190** | **0.00549** | **-0.0298** | **-0.00824** |
| sigmasq | 0.0249 | NA | NA | NA |

|  | Estimate | Std.Error | CI(lower) | CI(upper) |
| --- | --- | --- | --- | --- |
| (Intercept) | 1.464 | 0.3018 | 0.8729 | 2.056 |
| **Beta 1** | **0.531** | **0.0389** | **0.4552** | **0.608** |
| Alpha 52 | 0.111 | 0.0731 | -0.0328 | 0.254 |
| sigmasq | 0.012 | NA | NA | NA |

|  | Estimate | Std.Error | CI(lower) | CI(upper) |
| --- | --- | --- | --- | --- |
| (Intercept) | 1.19431 | 0.26365 | 0.6776 | 1.71e+00 |
| **Beta 1** | **0.53869** | **0.03975** | **0.4608** | **6.17e-01** |
| **Alpha 52** | **0.15678** | **0.07147** | **0.0167** | **2.97e-01** |
| **Linear Trend** | **-0.00632** | **0.00321** | **-0.0126** | **-3.45e-05** |
| sigmasq | 0.02117 | NA | NA | NA |

|  | Estimate | Std.Error | CI(lower) | CI(upper) |
| --- | --- | --- | --- | --- |
| (Intercept) | 1.8016 | 0.1707 | 1.467 | 2.136 |
| **Beta 1** | **0.6282** | **0.0351** | **0.559** | **0.697** |
| sigmasq | 0.0109 | NA | NA | NA |

Link function: log
Distribution family: nbinom (with overdispersion coefficient 'sigmasq')
Log-likelihood: -1940.87
AIC: 3887.74
BIC: 3900.081
QIC: 4082.899

